# A Similarity Metric, Rubric and Unified Hierarchy for Biomedical Publication Types and Study Designs

**DOI:** 10.64898/2026.01.03.26343378

**Authors:** Neil R. Smalheiser, Joe D. Menke, Arthur W. Holt

## Abstract

**Objective:** Our goal is to unify the 72 biomedical publication types and study designs (collectively, PTs) into a single rubric and hierarchy.

**Materials and Methods:** This is carried out in a data-driven manner by computing pairwise similarities of each PT against all others to form a similarity matrix. By performing hierarchical clustering we place each PT in a specific category and collect these into broader categories.

**Results:** Spearman correlations among PT pairs ranged from strongly negative to strongly positive (-0.732 to +0.997), with a mean of 0.176. Overall, we obtained 13 clusters of PTs and 5 more general categories: Observational Clinical Research, Qualitative and Genetic Methods, Clinical Evaluation and Validation, Interventional Trial Research, and Scholarly Synthesis and Discourse. These were then utilized to construct a unified hierarchy of PT terms.

**Discussion:** The rubric provides a flexible classification scheme for publication types and study designs that can accommodate new PTs as they are added over time.

**Conclusion:** The similarity metric has the potential to improve the modeling, implementation and evaluation of automated indexing systems. The PT rubric provides an overview that complements the existing NIH MeSH Hierarchy trees, and the unified hierarchy permits proper automated expansion for PT indexing and PubMed user queries involving PT terms.

## Background and Significance

Arguably the best curated indexing scheme for biomedical articles is the Medical Subject Heading (MeSH) terminology developed at the National Library of Medicine (NLM) (1). Each article in MEDLINE is annotated with a set of 8-20 terms, according to the most important topics discussed. The terminology is arranged first in categories and then placed in a set of hierarchical trees, according to their logical relationships. One of the trees deals with Publication Types such as Review, Letter, or Editorial (https://www.nlm.nih.gov/mesh/pubtypes.html), and others include study design-related terms as well as topical terms. Until recently all indexing was performed by PhD-level curators who read the full-text of each article and assign the most specific applicable term(s) on the hierarchy (2). Currently, however, most indexing is carried out by automated machine learning systems (3). The scheme is designed to facilitate high-recall retrieval of relevant articles when a user enters a free-text search query into the PubMed search interface. In the PubMed back-end, queries are processed to identify relevant MeSH terms and searches are automatically expanded to include synonyms and more specific terms that lie under them in the hierarchy (4).

In contrast to indexing of MeSH headings for the most important topics discussed in an article, some unique issues relate to automated indexing of articles in terms of publication types and study designs (for example, case-control studies, cross-over studies, etc.) (5–8). Our research team has been engaged in a long-term project along these lines (9–13). We seek to tag each article explicitly with all indexing terms that apply to them. In addition, we recognize that some articles are atypical; thus, we do not simply generate binary yes/no predictions but also assign probabilistic scores for each term. Together, this strategy is designed to ensure that a user seeking to retrieve all articles of a given study design can identify them with high precision and recall, and further, can adjust the threshold of the probabilistic scores to either maximize precision or recall as desired (14, 15). For simplicity, both publication types and study design-related terms will be collectively referred to as PTs.

During the course of our indexing project, we have identified several limitations regarding the NIH MeSH Hierarchy. First, the Publication Types form a separate tree apart from the study designs that are listed on multiple MeSH trees. Yet many Publication Types imply particular study designs; for example, Randomized Controlled Trial is listed as a Publication Type but has definite design features (random allocation, comparing an experimental group against a control group, generally interventional, and often double-blind). Thus, it would be desirable to merge publication types and study design-related MeSH terms into a single rubric. Second, the relative position of two terms on the hierarchical trees do not necessarily provide a measure of how similar they are. A similarity metric would provide a fuller understanding of how PTs relate to each other.

## Objective

Our goal is to unify the multiple trees containing publication types and study designs into a single rubric and hierarchy. This is carried out in a data-driven manner by computing pairwise similarities of each PT against all others to form a similarity matrix. By performing hierarchical clustering we place each PT in a specific category and collect these into broader categories.

## Materials and Methods

In a previous publication type indexing study (11) we processed the title and abstract of each article in the biomedical literature to create a vector representation of the article, then collected the vector representations of all articles belonging to a given PT and designated the centroid vector to represent the PT as a whole. In such a scheme, the similarity of two PTs can be measured in a straightforward manner as the distance between their centroids (16). This metric was employed in a SVM-based predictive model of 50 PTs called Multi-Tagger (11). However, more recently, we have utilized a transformer-based model that employed BERT-based encoders and represents articles not only in terms of text but some non-textual features as well (e.g., journal name, number of authors, number of references) (12, 13), and infers a probability value (0<p<1) that a given article belongs to each of 72 PTs. This model was employed here since it is more comprehensive and has been shown to have higher overall predictive performance for indexing PTs than the previous SVM-based model (12).

### Correlation analysis

The model probabilities produced by the transformer model were employed to obtain a pairwise similarity metric across all PTs. Full details of the model, its training data, and its architecture are given in Supplemental File 1. Briefly, the transformer model uses WeighCon supervised contrastive learning, and title, abstract, and various metadata-based features, such as journal name, served as input into the model. Training examples were taken from PubMed records published between 1987 and 2023 that had been manually assigned PTs by NLM, adjusting the number of examples per PT by under-sampling to deal with the fact that certain PTs have many more articles than others (12). In addition, using this same transformer model architecture, we added a new PT “Case Series”, designated from a curated corpus (17), combined with the data for the other PTs. Overall, this model was developed using a stratified random sample of 1,284,378 articles, which was stratified by PT to preserve label distribution. The dataset was split using 70/10/20 train/validation/test splits, again stratified to preserve label distribution in each split. Articles were tagged if the probabilistic prediction of the model was above a threshold, which was determined empirically to optimize for F1 within the validation set. The test set of articles from the modeling work, containing 256,897 articles, formed the basis for the present analysis and was further filtered to include only articles with complete abstracts, leaving 210,641 articles for analysis. (Articles lacking abstracts were excluded because they have limited textual features and on average show lesser predictive accuracy (12).)

From the model-predicted probabilities for each of the 72 publication types across the articles of the test set, a nonlinear (Spearman) correlation rho was computed for all 2,556 pairwise combinations of the 72 publication types (illustrated in Figure 1) to form a 72x72 correlation matrix (Supplemental File 2).

**Figure 1.**
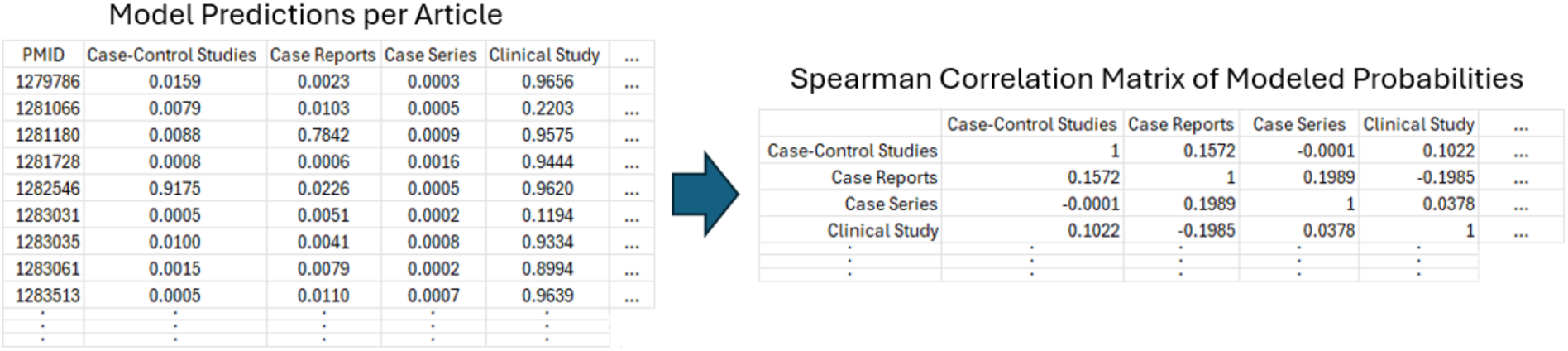
Correlation Matrix Derivation. Modeled predicted probabilities were used to compute Spearman correlations for each pairwise combination of publication types.

## Results

### Distribution of PT Similarity Measures

Model score correlations among PT pairs ranged from strongly negative to strongly positive (-0.732 to +0.997), with a mean of 0.176. The distribution appears to be bimodal with one mode centered near zero correlation, and a shoulder centered around 0.59 (Figure 2). The highest positive correlations relate to clinical trial subtypes and related clinical study methodologies. The negative correlations consist of publication type pairs that are functionally unrelated, such as Clinical Study vs. Biography.

**Figure 2.**
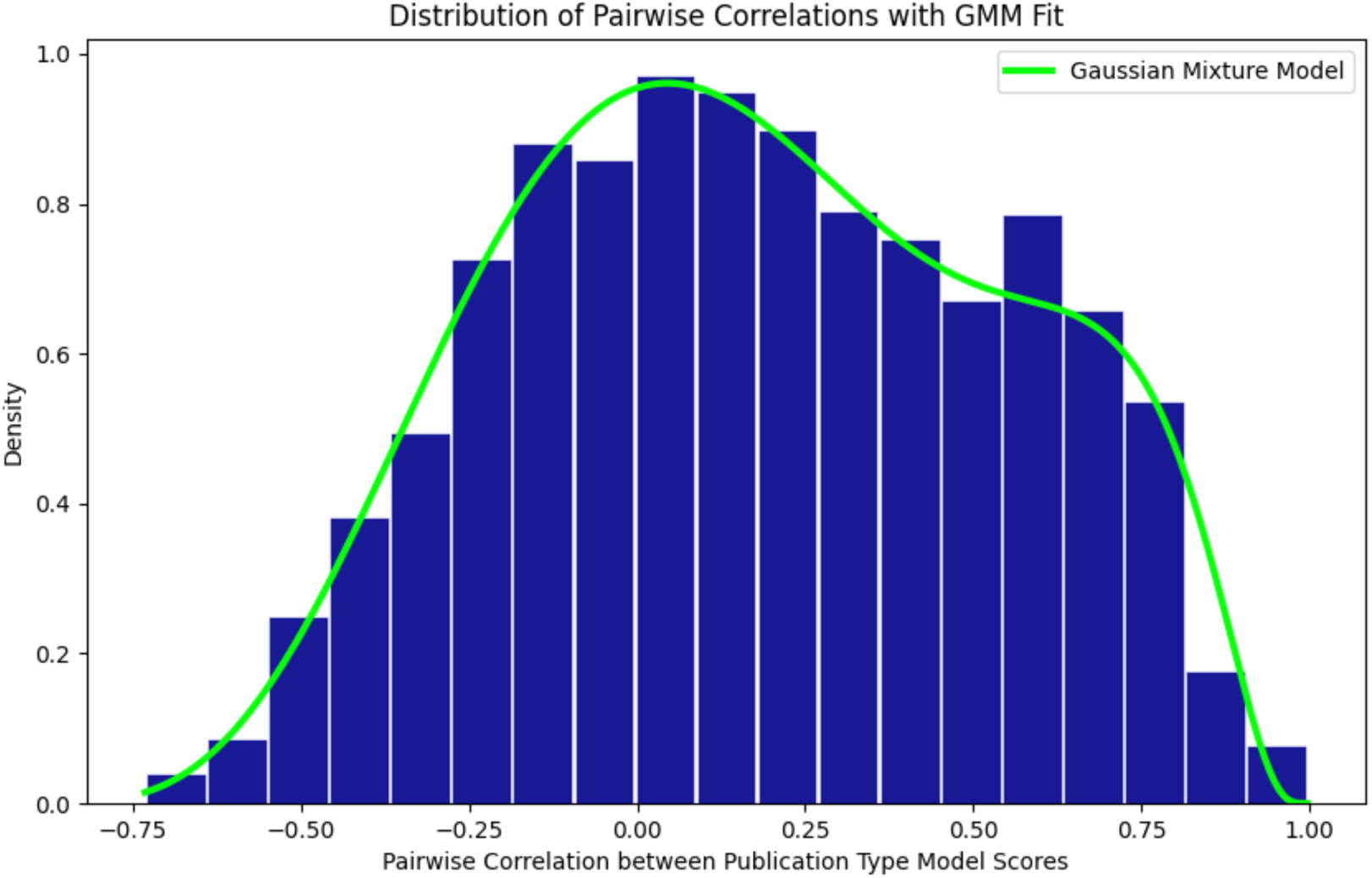
Distribution of Pairwise Correlations for PTs. Spearman correlation was computed among model-predicted probabilities for assignment of 72 publication types forming a 72x72 correlation matrix. The distribution of the correlation coefficients is bimodal with one mode centered around zero, the other around 0.57.

### Hierarchical clustering of publication types and study designs

Several different clustering techniques were evaluated, including graph-based community detection methods and statistical clustering methods. The graph-based clustering methods produced reasonable communities, but were found to be sensitive to noise and joined disparate publication types such as case reports with editorials and commentary. Agglomerative hierarchical clustering provided a more reasonable and functional basis for grouping PTs using the pairwise correlation matrix inverted to represent distances and joined with Ward linkage (Figure 3) (18).

**Figure 3.**
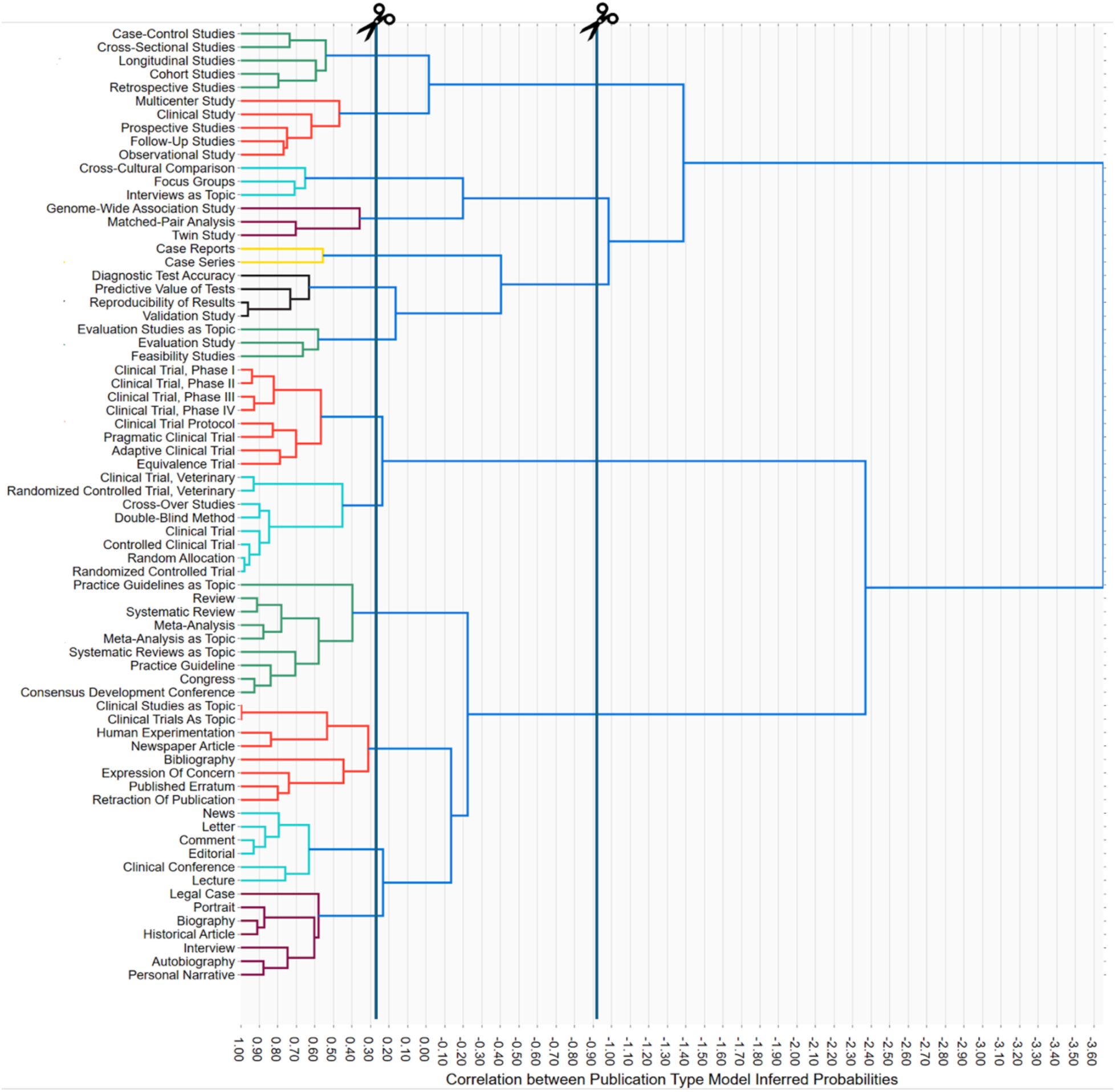
Hierarchical Clustering of Publication Types by Model-Inferred Probabilities. Publication type clusters derived from pairwise model score correlations and grouped with hierarchical clustering. Cut points were chosen to delineate 13 low-level categories and 5 broader categories.

Starting with the most similar pairs of PTs, several different categories naturally emerged in a data-driven manner (Figure 3). For example, the category of Observational Epidemiologic Study Designs comprised case-control, cross-sectional, longitudinal, cohort and retrospective studies. Interestingly, other types of observational studies fell into a distinct category, General Clinical and Observational Studies, consisting of both publication types and MeSH study designs, namely, multicenter study, clinical study, prospective studies, follow-up studies and observational study.

Note that the clustering scheme reflects the behavior of NLM indexers, who attach only the most specific indexing term that applies to a given article. Thus, even though the Clinical Study PT definition comprises both interventional and observational designs (https://www.ncbi.nlm.nih.gov/mesh/2009830), if the article is interventional, NLM indexers would affix the more specific term Clinical Trial instead. For this reason, in Figure 3, Clinical Study is placed with observational designs, whereas Clinical Trial (logically a subset of Clinical Study) is placed with interventional designs. Similarly, if an observational study had, say, a cohort design, it would be indexed as Cohort Studies instead of Observational Study, explaining why the two PTs were placed in distinct clusters. The relationships shown in Figure 3 are thus complementary to the hierarchical relationships and trees displayed in the NIH Hierarchies.

Overall, we obtained 13 low-level clusters of PTs and 5 more general categories: Observational Clinical Research, Qualitative and Genetic Methods, Clinical Evaluation and Validation, Interventional Trial Research, and Scholarly Synthesis and Discourse. One PT, Scientific Integrity Review, was excluded from the clustering process because very few articles in our dataset were tagged with that type; due to its nature (consisting of official scientific misconduct findings), it was manually placed in Scholarly Publishing and Research Integrity. Table 1 contains a full list of publication types and study designs examined along with category assignments. Another view is presented in Supplemental File 2.

**Table 1.**
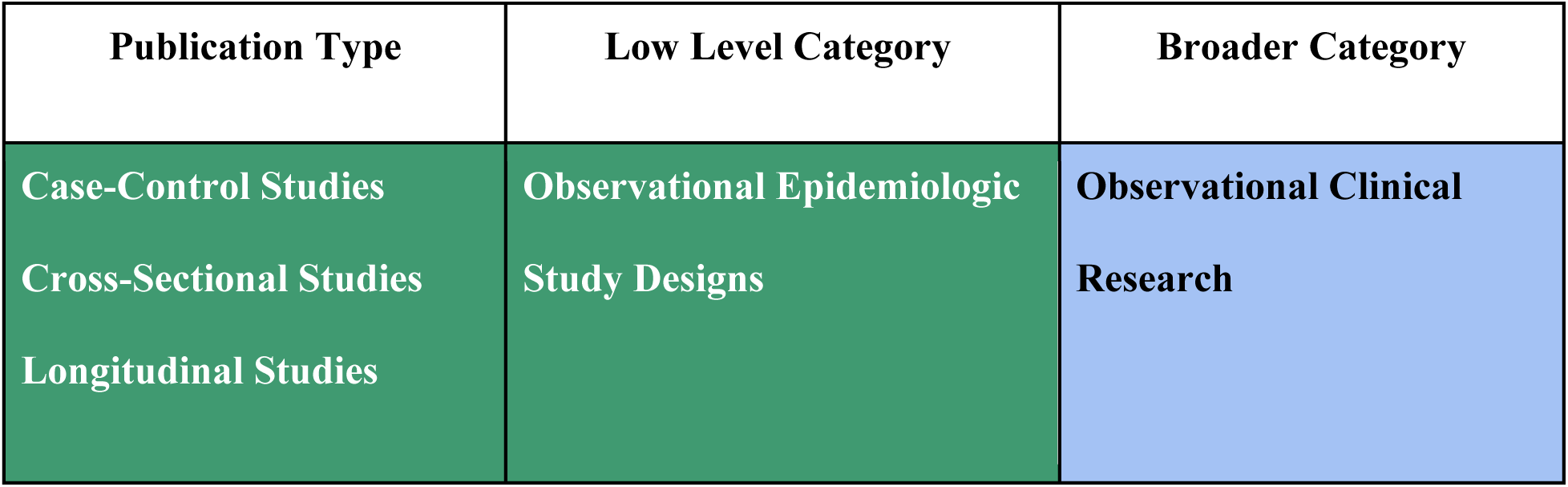
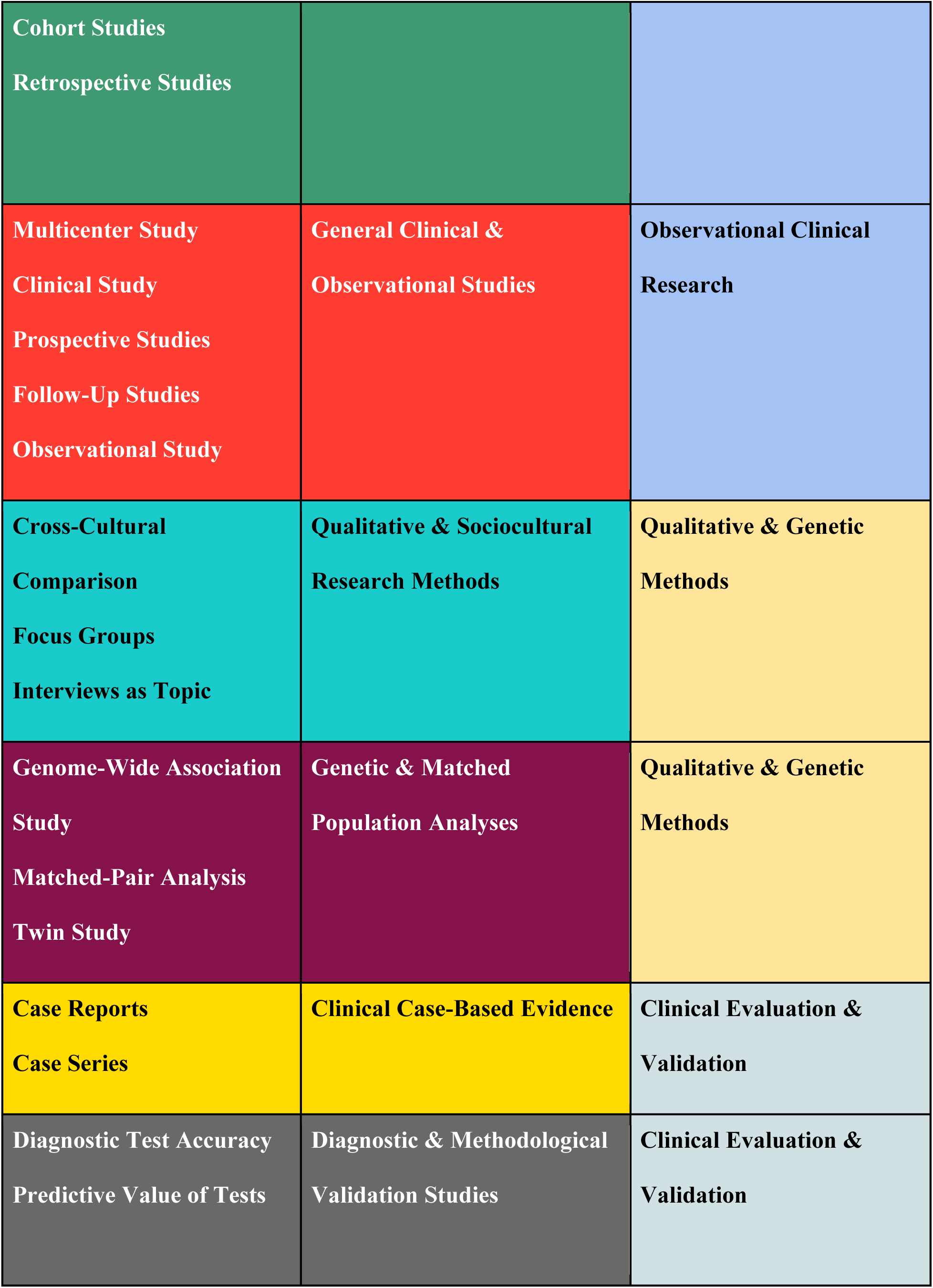
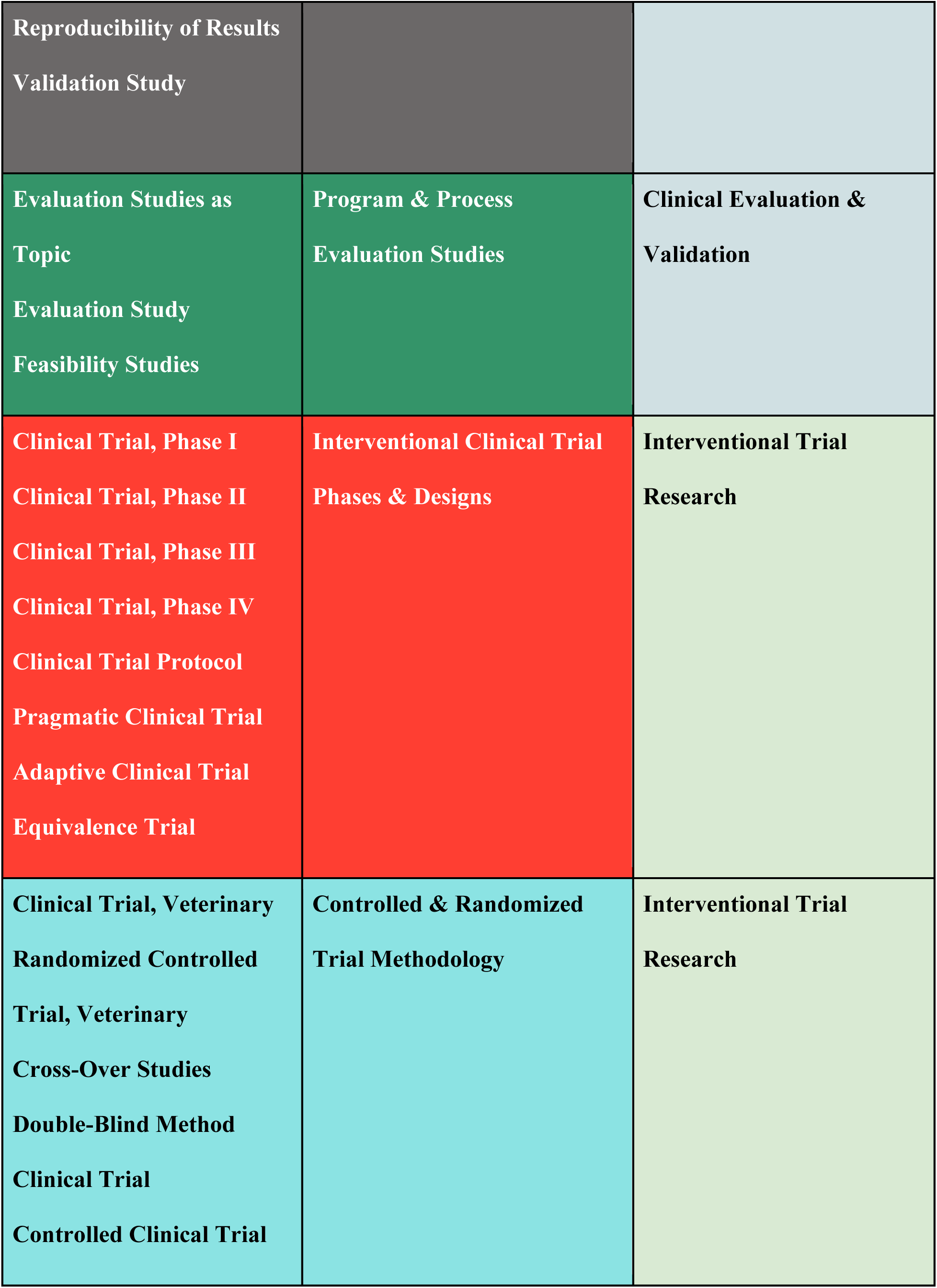
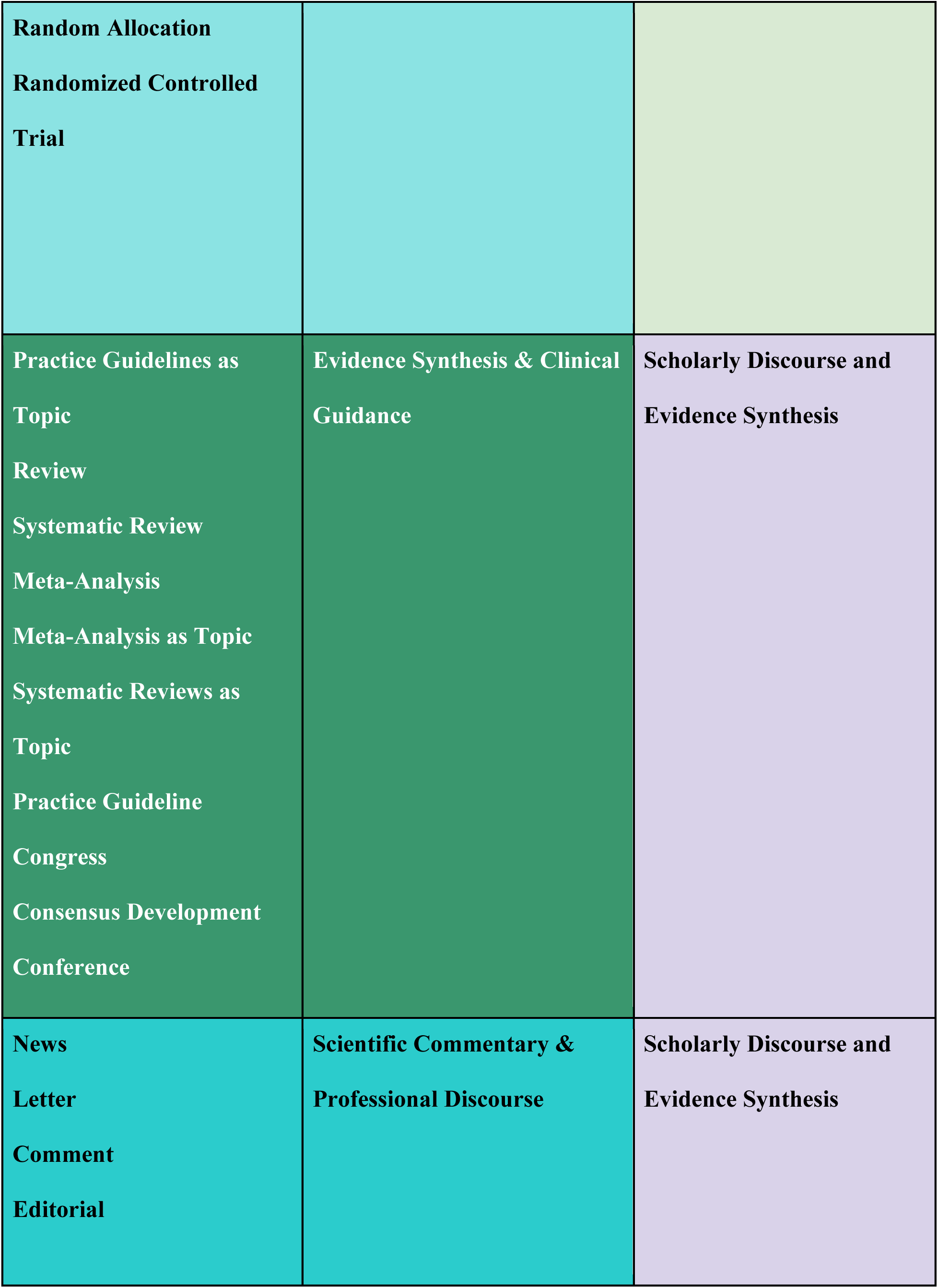
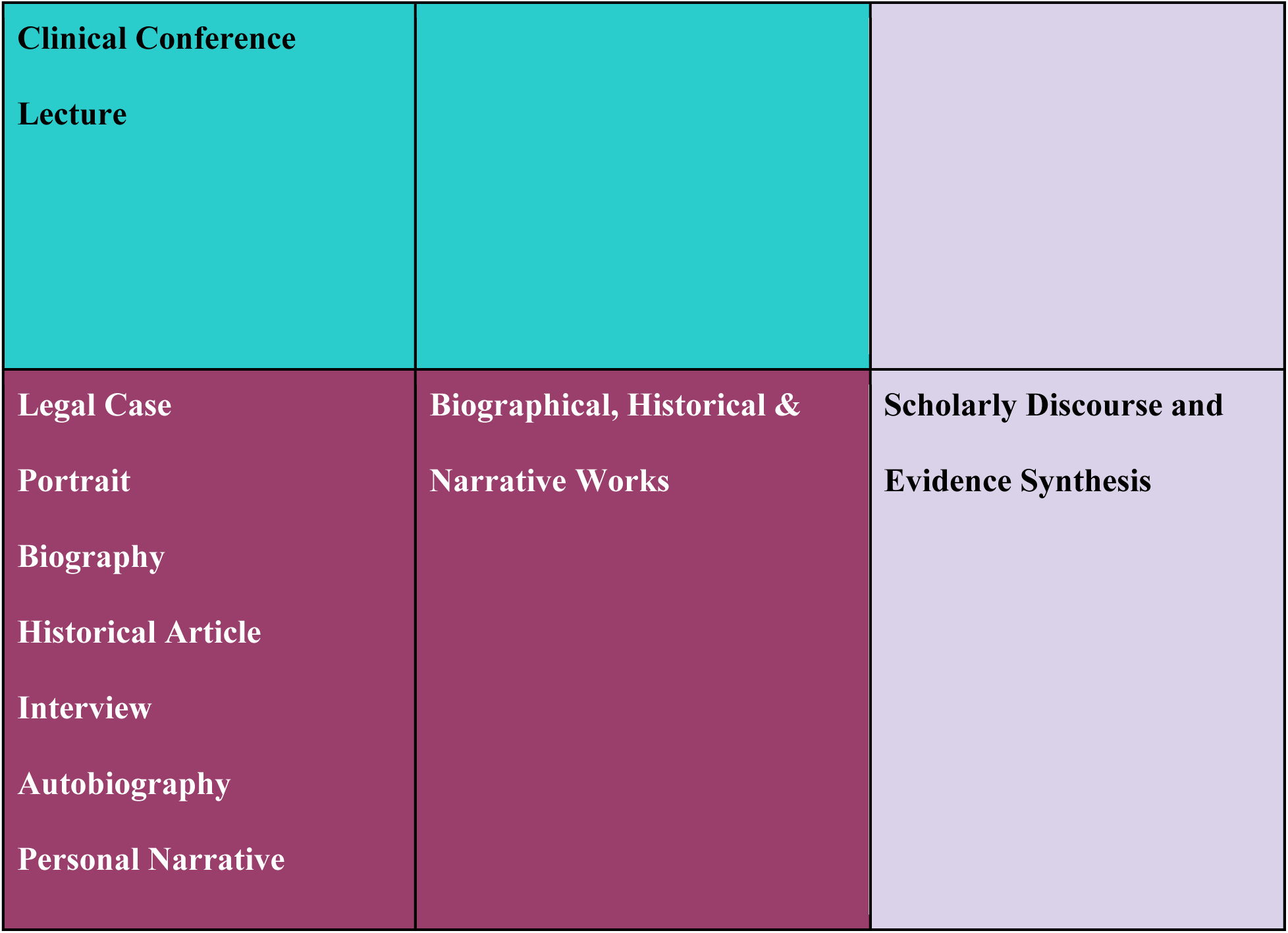
Publication Types and Observed Cluster Assignments.

### A unified hierarchy of publication types and study designs

The rubric presented so far merges PT terms that reside across multiple trees in the NLM MeSH hierarchy. However, it still remains to organize the terms into a single hierarchy that reflects their logical relationships, which is necessary in order to perform proper automated expansion for PT indexing and retrieval of user queries. We constructed the hierarchy manually, being informed by three sources of evidence: a) maintaining the existing MeSH Hierarchy placements whenever possible, b) considering the nearest-neighbor PTs and rubric categories (Figure 3 and Table 1), and c) examining the automated query expansion of terms as performed by PubMed. The proposed unified hierarchy is shown in Table 2.

**Table 2.**
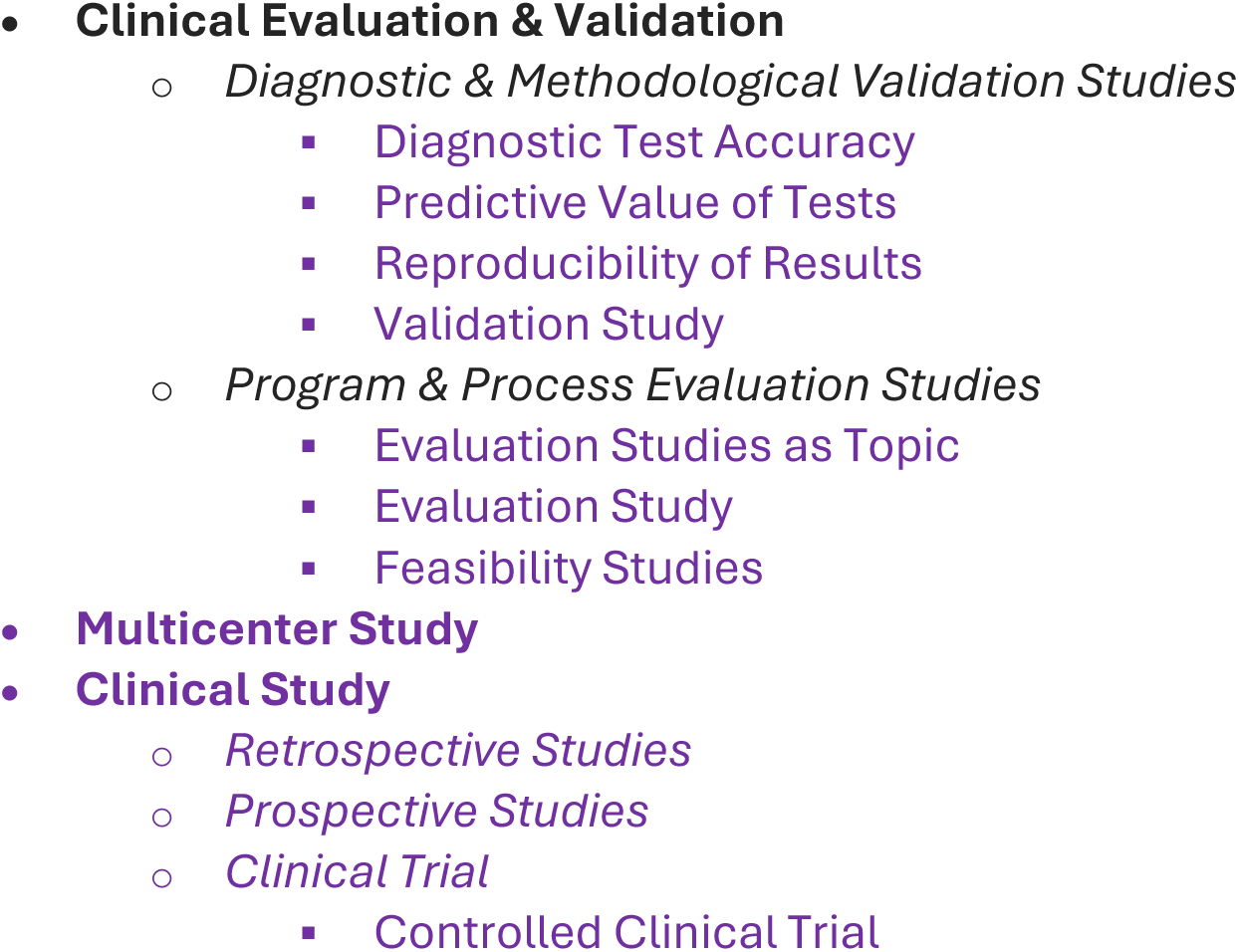
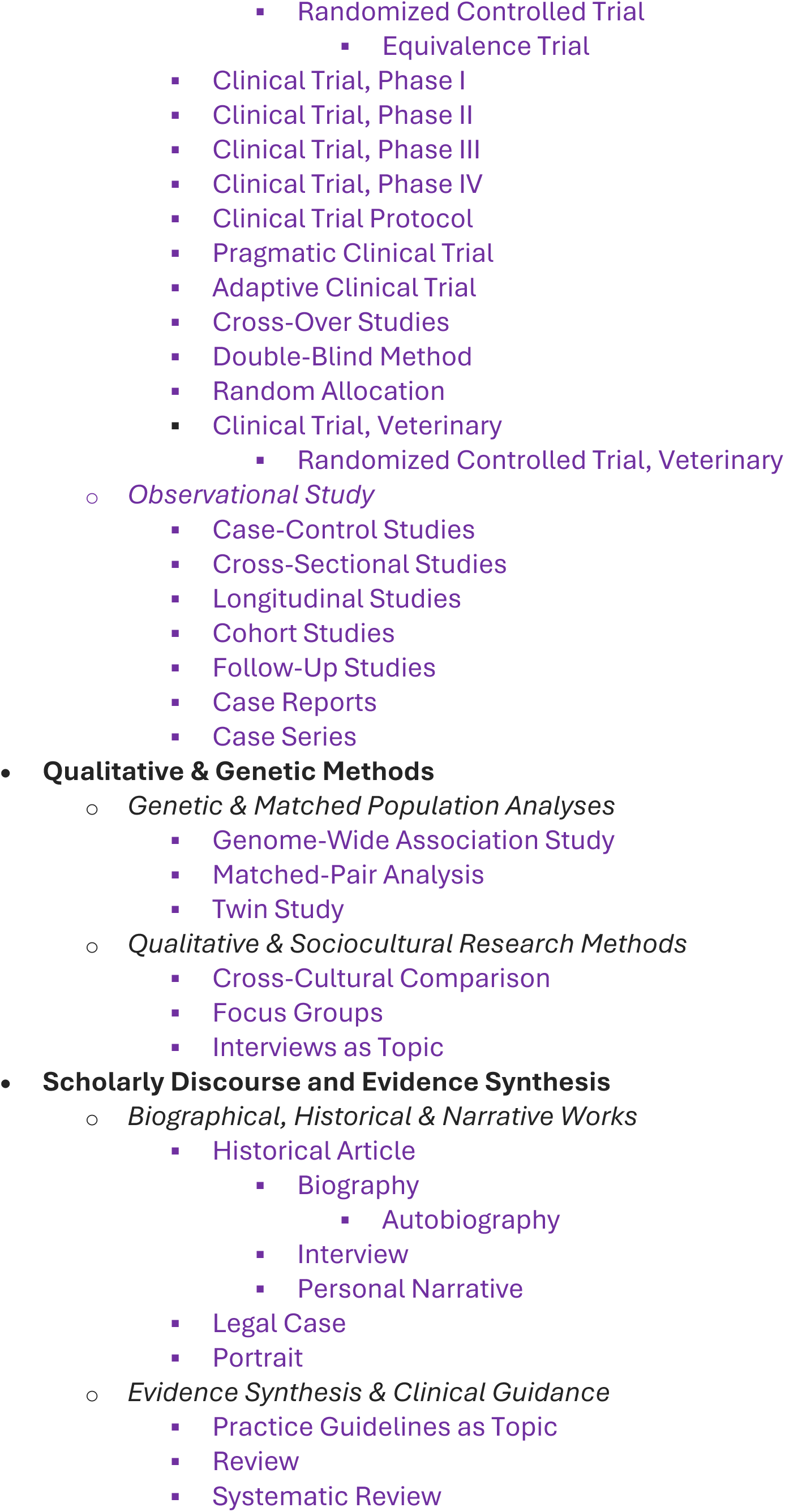
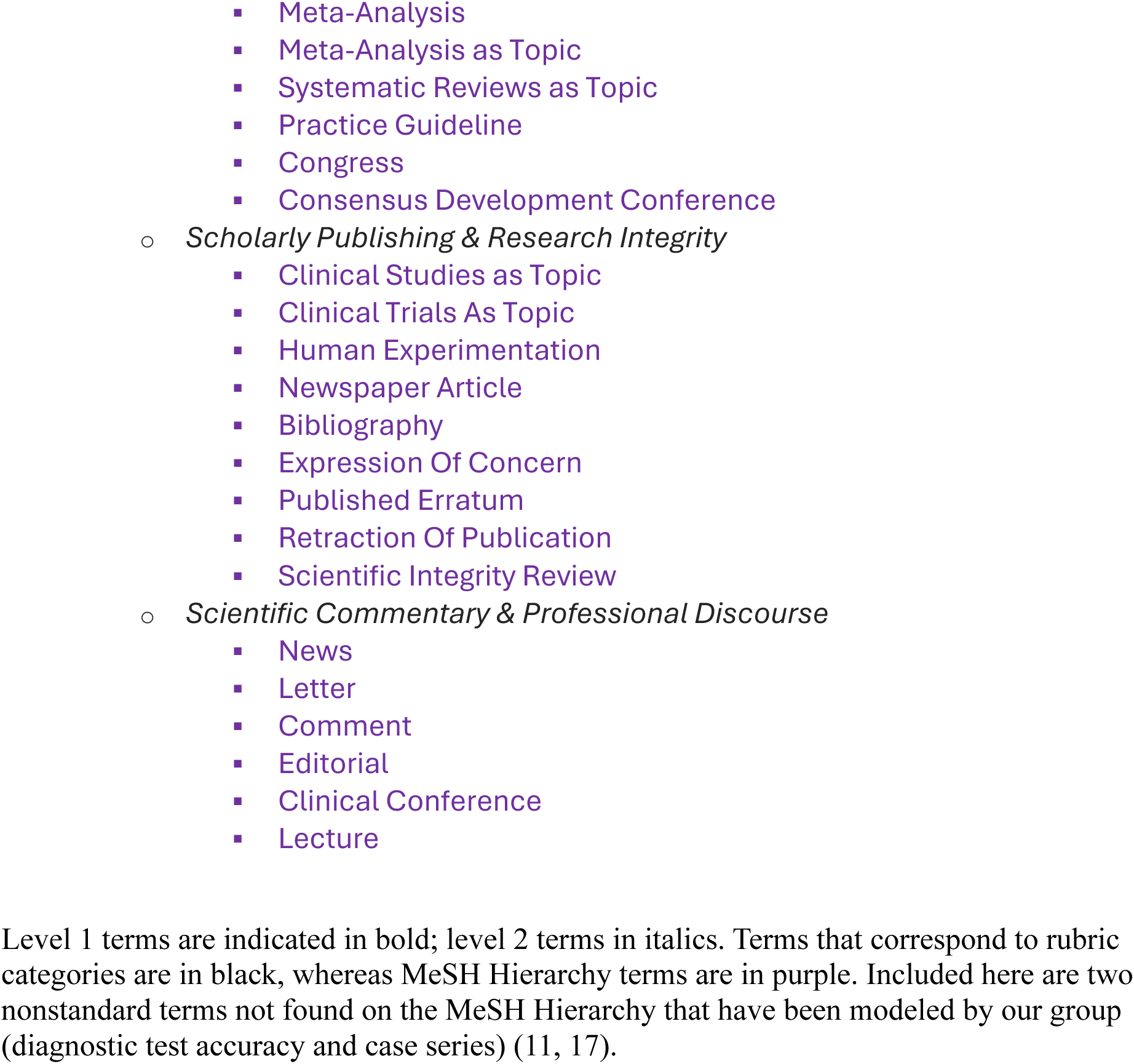
Unified Hierarchy of Publication Types and Study Designs.

The placement of several PT terms in Table 2 deserves notice. 1) “Clinical Trial Protocol” is indexed under “Clinical Study” in the MeSH Hierarchy. We kept this arrangement in our hierarchy, but note that protocol articles should ideally also be automatically indexed as “Clinical Trials as Topic” as well. 3) “Systematic Review” is placed under “Review” agreeing with the automatic expansion carried out by PubMed (though this differs from its placement in the MeSH Hierarchy). 4) All observational study designs are placed under “Observational Study”; this implies that all articles having observational designs (e.g., cohort studies) should automatically be assigned Observational Study as well. Note that this is not currently carried out by PubMed via automated query expansion. 5) “Retrospective Studies” and “Prospective Studies” are no longer placed under either “Cohort Studies” or “Case-Control Studies”, but are placed directly under Clinical Study. This corrects a major anomaly in the existing MeSH Hierarchy, since many articles with retrospective or prospective designs are actually neither cohort studies nor case-control studies. In fact, although the existing MeSH Hierarchy considers both Retrospective and Prospective as observational study designs, a minority (∼20%) of articles indexed by NLM as Retrospective or Prospective are also indexed as Clinical Trial. Because these study designs can be either observational or interventional, we place Retrospective and Prospective designs directly under Clinical Study.

## Discussion

The formulation of a similarity metric for PTs led to the construction of a unified rubric that provides a simple, concrete overview of the many biomedical publication types and study designs, in a single diagram. This provides a complementary view from the NIH hierarchies which identify different terms in terms of their logical relationships and proximity on several independent trees. Cid and Mork previously constructed a PT correlation matrix based on the frequency of co-occurrence of multiple PTs appearing on individual articles (8) but their scheme did not include study design-related MeSH terms and only considers articles that received multiple PT terms. In contrast, our similarity metric was computed across all articles and reflects the indexing decisions of NLM indexers, who generally attach only the most specific indexing term that applies to a given article. Hence our scheme provides a more comprehensive, finer-grained overview.

As the similarity metric is data-driven, it can be recalculated to adapt to future changes in publications, and can easily accommodate new indexing terms as they arise over time. For example, since NLM currently has no formal indexing of case series articles, we recently created a new publication type, Case Series (17). After adding this PT to the existing transformer model we obtained correlation values which placed Case Series next to Case Reports (Figure 2).

Apart from providing a classification scheme for publication types and study designs, we believe that the similarity metric should have utility for better automated PT indexing. With regard to the current transformer model, we use a one-against-all model in which each article is assessed for a given PT vs. all others. However, the training process should be sharpened if one used training examples of one PT as negative examples against another PT that is very similar (but not a subset of the other). For example, Case Reports and Case Series are nearest neighbor PTs in the rubric. Case reports tend to be incidental observations of one or a few patients, whereas case series tend to be planned observations of four or more patients, but the two types of article are not sharply delineated (17). By comparing and contrasting these two PTs during model training, the probabilistic scores should better discriminate whether a given article has typical case report-like vs. typical case series-like features.

The similarity metric may also assist in evaluating the errors made by an automated indexing system. Since the probabilistic predictive scores for nearest-neighbor PTs have high Spearman correlation values, one would expect that a mis-assigned article would most likely be tagged with the nearest neighbor PT instead. In many cases, the errors might be of little import (e.g., mis-assigning a Focus Group article as Interviews as Topic). Other nearest-neighbor errors have high importance for high recall retrieval, for example, mis-assigning Cohort Studies as Case-Control Studies. Again, identifying these cases would prioritize them for contrastive training in which one PT is used as negative examples for the other PT.

Another possible use for the rubric is as a framework for alternative LLM-based assignment of publication types and study designs (6). For example, the organization of PTs into small and larger categories allows one to envision a Twenty Questions procedure in which the LLM would first attempt to place an article into the most appropriate broad category (e.g. observational vs. interventional research) and then to ascertain the best specific category and the best choice among the PTs in that category.

This study has several limitations. The processing of articles was limited to articles containing abstracts, which provide more complete data for model predictive scores; this will under-sample a few PTs that have a high proportion of articles lacking abstracts (e.g., Letter or Portrait).

However, these PTs tend to be the least important in terms of clinical relevance and should have little effect on the rubric as a whole. Another limitation is that agglomerative hierarchical clustering’s optimal solution is not necessarily unique. However, although calculated nearest-neighbors can vary slightly across different test sets, different model parameters, and different examined time periods, the placement of PTs into categories is likely to be robust and stable over time.

Finally, the unified rubric informed the construction of a single PT hierarchy, that merges PT terms across multiple trees of the MeSH Hierarchy and corrects several of its deficiencies and anomalies (Table 2). The unified hierarchy permits better automated expansion for PT indexing; for example, all observational study designs are now automatically assigned Observational Study as well. We propose that this hierarchy is also preferred for PubMed user queries, for automated expansion of PT terms.

## Conclusion

We present here pairwise similarity measures that allowed us to construct a rubric that arranges publication types and study designs in a unified diagram and a unified hierarchy. Such a rubric complements the existing NIH hierarchies and has the potential to improve the modeling, implementation and evaluation of automated indexing systems.

## Data Availability

The nonlinear (Spearman) correlation rho was computed for all pairwise combinations of the 72 publication types (illustrated in Figure 1) to form a 72x72 correlation matrix. This is shown in Supplemental File 2.

## Author Contributions

AWH: Methodology, Formal analysis, Investigation, Writing – review & editing. JDM: Methodology, Software, Validation.

NRS: Conceptualization, Funding acquisition, Methodology, Supervision, Writing - original draft, Writing – review & editing.

---

**Supplemental File 1.** Description of the transformer model used to generate model probabilities for publication types and study designs.

**Supplemental File 2.** Publication type details, including low level and broader category assignments, are provided in an Excel workbook posted in the Dryad repository http://datadryad.org/share/LINK_NOT_FOR_PUBLICATION/2BTwJF2CCtcfHm4Ug_IYJ766irwfbBWwD1qN6owxFj8 containing two tabs as follows:

**Unified Rubric** - a table with 73 rows, including header row, with the following columns:

- **Publication Type** - names of publication types/study designs with capitalization designed to match NLM spelling
- **Low Level Category Number** - an integer assigned to each of the 13 low level categories described in Table 1
- **Display Sort Order** - an integer indicating position in the dendrogram presented in Figure 2
- **Low Level Category** - descriptive name for the 13 categories to which the PT is assigned
- **Broader Category** - descriptive name for the 5 broader categories to which the PT is assigned
- **Top Correlated PT to 5th Correlated PT** - the top 5 most correlated PTs for each PT, along with Spearman correlation values
- **Least Correlated PT & Least Correlation** - the PT having the least (most negative) correlation value

**Correlation Matrix** - a 72x72 square matrix containing the computed pairwise correlations among the 72 PTs examined (as in Figure 1)

