## Supplemental File 1 for "A Similarity Metric, Rubric and Unified Hierarchy for Biomedical Publication Types and Study Designs" (from another version of this preprint: version 1, 2026-01-04)

**Supplemental File 1. Description of the transformer model used to generate model probabilities for publication types and study designs.** The model was developed with 1,284,986 PubMed articles, split into training, validation, and test sets using a 70/10/20 ratio (899,819 / 128,303 / 256,864). Articles were sampled and split using stratified random sampling to preserve the multi-label publication type (PT) distribution observed in PubMed as a whole. Articles were eligible for inclusion if they were published between 1987 and 2023 in English or had an English abstract. No additional exclusion criteria were applied (e.g., articles without abstracts were still included for training; they were represented using title and metadata features where available).

To mitigate class imbalance between common and rare PTs, we applied undersampling to the training set, resulting in 702,603 training articles; validation and test sets were left unchanged. The undersampling procedure is described more thoroughly in our prior work [12]. Input features followed the configuration described in prior work [13] and, when all features were available, consisted of the title, abstract, and verbalized metadata fields (e.g., journal, number of authors, etc.), concatenated and tokenized using the SPECTER2 tokenizer with a maximum sequence length of 512 tokens. Any inputs longer than 512 tokens were truncated.

The model architecture consisted of a SPECTER2-base encoder and a linear classification head with sigmoid activation, producing multi-label predictions for 72 publication types and study designs.[19] Training used asymmetric loss with parameters clip = 0, γ_neg = 0, and γ_pos = 0, equivalent to binary cross-entropy under these settings, with label smoothing (α = 0.05). In addition to the classification objective, training incorporated both unsupervised and supervised contrastive learning: ADNCE loss for unsupervised contrastive learning using dropout-based augmentation (dropout = 0.1) with weights w₁ = 1.0 and w₂ = 1.0, and HeroCon loss for supervised contrastive learning using articles sharing at least one PT label as positive anchors. The total training loss was computed as a weighted combination of the classification and contrastive objectives: loss = mean (α L_unsup  + β * L_sup + L_asl) where α = 0.01 and β = 0.1.

Models were trained for up to 15 epochs using the RAdam optimizer with a learning rate of 1e-5 for SPECTER2 layers and 1e-2 for the classification head, a batch size of 32, and mixed-precision training enabled.[20]  Early stopping was applied based on macro-F1 performance on the validation set with training stopped if there was no improvement for three consecutive epochs. Binary decision thresholds were tuned independently for each label on the validation set to maximize F1 score and were fixed for final evaluation on the test set. Training and inference were performed on a single A100 32GB GPU using Python 3.9.18 and PyTorch 2.2.0 built for CUDA 12.1.
